# Detecting Problematic Opioid Use in the Electronic Health Record: Automation of the Addiction Behaviors Checklist in a Chronic Pain Population

**DOI:** 10.1101/2023.06.08.23290894

**Authors:** Angus H. Chatham, Eli D. Bradley, Lori Schirle, Sandra Sanchez-Roige, David C. Samuels, Alvin D. Jeffery

**Affiliations:** Vanderbilt University School of Nursing, Nashville, TN, USA; Department of Anesthesiology, School of Medicine, Vanderbilt University, Nashville, TN, USA; Department of Psychiatry, University of California San Diego, La Jolla, CA, USA; Department of Medicine, Division of Genetic Medicine, Vanderbilt University Medical Center, Nashville, TN, USA; Department of Molecular Physiology and Biophysics, Vanderbilt Genetics Institute, Vanderbilt University, Nashville, TN, USA; Department of Biomedical Informatics, Vanderbilt University Medical Center, Nashville, TN, USA

## Abstract

**Importance:** Individuals whose chronic pain is managed with opioids are at high risk of developing an opioid use disorder. Large data sets, such as electronic health records, are required for conducting studies that assist with identification and management of problematic opioid use.

**Objective:** Determine whether regular expressions, a highly interpretable natural language processing technique, could automate a validated clinical tool (Addiction Behaviors Checklist^1^) to expedite the identification of problematic opioid use in the electronic health record.

**Design:** This cross-sectional study reports on a retrospective cohort with data analyzed from 2021 through 2023. The approach was evaluated against a blinded, manually reviewed holdout test set of 100 patients.

**Setting:** The study used data from Vanderbilt University Medical Center’s Synthetic Derivative, a de-identified version of the electronic health record for research purposes.

**Participants:** This cohort comprised 8,063 individuals with chronic pain. Chronic pain was defined by International Classification of Disease codes occurring on at least two different days.^18^ We collected demographic, billing code, and free-text notes from patients’ electronic health records.

**Main Outcomes and Measures:** The primary outcome was the evaluation of the automated method in identifying patients demonstrating problematic opioid use and its comparison to opioid use disorder diagnostic codes. We evaluated the methods with F1 scores and areas under the curve - indicators of sensitivity, specificity, and positive and negative predictive value.

**Results:** The cohort comprised 8,063 individuals with chronic pain (mean [SD] age at earliest chronic pain diagnosis, 56.2 [16.3] years; 5081 [63.0%] females; 2982 [37.0%] male patients; 76 [1.0%] Asian, 1336 [16.6%] Black, 56 [1.0%] other, 30 [0.4%] unknown race patients, and 6499 [80.6%] White; 135 [1.7%] Hispanic/Latino, 7898 [98.0%] Non-Hispanic/Latino, and 30 [0.4%] unknown ethnicity patients). The automated approach identified individuals with problematic opioid use that were missed by diagnostic codes and outperformed diagnostic codes in F1 scores (0.74 vs. 0.08) and areas under the curve (0.82 vs 0.52).

**Conclusions and Relevance:** This automated data extraction technique can facilitate earlier identification of people at-risk for, and suffering from, problematic opioid use, and create new opportunities for studying long-term sequelae of opioid pain management.

**Key Points:** *Question:* Can an interpretable natural language processing method automate a valid, reliable clinical tool in order to expedite the identification of problematic opioid use in the electronic health record?

*Findings:* In this cross-sectional study of patients with chronic pain, an automated natural language processing approach identified individuals with problematic opioid use that were missed by diagnostic codes.

*Meaning:* Regular expressions can be used in automatically identifying problematic opioid use in an interpretable and generalizable manner.

## Introduction

Chronic pain affects more than 40 million individuals in the United States, of which approximately 10 million experience high-impact chronic pain affecting daily activities.^2^ Prescription opioids are a primary treatment for chronic pain management. Given the highly addictive nature of opioids, the risk of developing an opioid use disorder (OUD) is estimated to be high at approximately 18%.^3^ OUD is associated with a financial burden of more than $1 trillion when accounting for health care, lost work productivity, and criminal justice costs.^4^ In order to address this problem from a healthcare perspective, we must first be able to identify which patients suffer from OUD and/or are at risk for developing OUD.

The magnitude of this particular problem necessitates large-scale data sources for identifying individuals across the continuum of problematic opioid use. Currently, the largest source of health data is the electronic health record (EHR) used for routine clinical care. The standard method for detecting a clinical problem in the EHR is through diagnostic indicators, such as problem lists or International Classification of Diseases (ICD) codes used for billing purposes.^5,6^ However, ICD codes are not a reliable source of OUD diagnosis because the codes are under-utilized, which has been attributed to OUD-related stigma and provider concerns about barriers to future pain management.^3,7–10^

Expanding the search to additional areas that contain clinical notes has been explored. For example, Palmer et al. used natural language processing (NLP) based on matching terms in a customized dictionary of 1,248 problematic opioid use keywords developed by subject matter experts. They discovered NLP techniques could identify many individuals with problematic opioid use that did not have relevant ICD codes; however, they also found many patients with relevant ICD codes who were not flagged by NLP methods.^11^ Similar results were reported by Carrell et al. who developed a customized dictionary (of 1,288 unique terms) based on recommendations from subject matter experts and iterative reviews of example text.^12^ The results from these papers would suggest limitations to a customized dictionary approach to NLP (or an inadequacy of EHR documentation).

The Addiction Behaviors Checklist (ABC) is a valid and reliable instrument that can be used for identifying OUD risk among chronic pain patients.^1^ The ABC collects risk information provided by clinicians, making it a particularly suitable tool to adapt to EHR data. We used the ABC instrument instead of other OUD risk assessment tools (e.g., the Revised Opioid Risk Tool^13^) because the ABC collects risk information from language patterns used by clinicians, who are the authors of the notes used in this study. Use of such an assessment tool could guide the NLP methods for automatically searching the clinicians’ notes within the EHR. NLP can be leveraged for automating information extraction from text documents and includes techniques ranging from pattern matching to concept extraction to advanced numerical vector embeddings.^14^ If an NLP method that is highly interpretable by clinicians, such as one that facilitates review of matching text, could perform comparably to manual chart reviews, there is potential to expedite and scale the identification of addictive behaviors within EHRs.

Therefore, the objective of this study was to leverage an interpretable NLP technique to automate the ABC instrument for purposes of expediting research or clinical chart reviews.

## Materials and Methods

### Cohort Definition and Data Collection

We conducted a retrospective, observational cohort study of individuals with chronic pain. We selected this phenotype because individuals with chronic pain are known to have a higher incidence of opioid use and OUD than the general population.^15^ We studied this cohort in our prior publication.^16^ We derived the study data from Vanderbilt University Medical Center’s Synthetic Derivative, a de-identified version of the electronic health record for research purposes.^17^ Chronic pain was defined based on ICD codes (e.g., 338.2, 338.21, G89.2, G89.21 – see Appendix A for a full list of ICD codes) occurring on at least two different days to decrease false positives when including patients with only a spurious code.^18^ We collected all electronic health record free-text notes (e.g., progress notes, patient communication, history and physical) for a patient restricted to the window 30 days before the patient’s first chronic pain ICD code through 30 days after the last chronic pain ICD code. We also collected demographic information (age, gender, ethnicity, and race) from the electronic health record to characterize our population. We limited the cohort to individuals aged 13 years or older at time of their first chronic pain diagnosis.

### Instrument Development

We used regular expressions to identify text patterns in the clinical notes. Regular expressions are an easily implementable and interpretable form of NLP that can be used to search for pattern matches in a corpus of text.^14^ We used the ABC ^1^ to guide regular expression development. For each item in the checklist, three members of the research team (a health sciences undergraduate student [Chatham], a biomedical data scientist [Bradley], and a PhD nurse practitioner and informatician [Jeffery]) generated one or more regular expressions to operationalize the conceptual intention of the item. After drafting a regular expression, two separate members of the research team (a pain and opioid researcher [Schirle] and a substance use disorder geneticist [Sanchez-Roige]) manually reviewed performance of the candidate expression by examining 50-100 positive matches in the training data set.

Following a review of matches, we examined whether additional filtering for matches near 133 opioid-related terms (Appendix B) and/or at least 7 negation detection terms (Appendix C) improved performance. For example, with ABC item 2 (“Patient has hoarded meds.”), we used regular expressions to search for “hoard” and then filtered to include only those variations of “hoard”, “stash”, “left over”, “storing”, and “stockpil”[sic] that were followed by variations of “pain med,” “opioid”, “opiod”[sic], “narc”, “analges”[sic] or an opioid drug name. Then, of those sentences, any that included a negating term preceding “hoard” (or one of the related verbs) were excluded. We added a final step for some expressions where we included common false positive matches. For example, opioids were frequently mentioned in the Discharge Instructions of a patient’s chart. We removed opioid matches if they were preceded by the phrase “Discharge Instructions.”

After a candidate regular expression’s matches were reviewed in the training data set, modifications were made to expressions based on suggestions from Schirle and Sanchez-Roige. New examples of matches were generated by Chatham, Bradley, and Jeffery in another group of 50-100 randomly selected matches. The iterative process of regular expression development resulted in 27 regular expressions (see Appendix D) representing the ABC items. We implemented the regular expressions in python (version 3.10). We applied each regular expression to every clinical note. If 1 or more matches for a given ABC item were discovered in any of the patient’s notes within the time window that defined the cohort, that patient received 1 point toward an overall total score.

### Statistical Analysis

We evaluated our methods against a gold-standard manual review in a holdout test set of 100 patients that had been adjudicated in our previous study^16^ where we classified individuals as having no, some, or high evidence of OUD and substance use disorders (SUD). These 100 patients were randomly selected from the chronic pain cohort and were only used for evaluating our phenotyping methods. We reviewed patients’ records guided by a keyword template based on the Diagnostic and Statistical Manual of Mental Disorders, 5^th^ Ed. (DSM V) criteria for OUD,^19^ the ABC instrument,^1^ and others’ studies focused on detecting problematic opioid use within EHR data.^11,12,20^

We calculated the sensitivity (i.e., recall - proportion of cases with a match), specificity (proportion of controls without a match), precision (i.e., positive predictive value [PPV] - proportion of matches that were cases), negative predictive value ([NPV] proportion of non-matches that were controls), and F1-score (harmonic mean of sensitivity and positive predictive value) of our regular expression scoring system against the manually adjudicated labels (n=100). We examined the pairwise phi coefficients (for binary variables) between each item of the ABC instrument in the entire dataset (n=8,063). We compared the presence of OUD and SUD ICD codes (see Appendix A for full list) present in individuals’ records to serve as another source of validation. We used recall-precision curves and area under the receiver operating characteristic curves (AUC) to evaluate performance of both the total ABC score and the OUD ICD codes against the manual reviews.

This study follows the Strengthening the Reporting of Observational Studies in Epidemiology (STROBE) reporting guidelines.^21^ We acquired ethics approval under Institutional Review Board study #181443 and #201918. All code is publicly available at [unblinded URL after review].

## Results

### Cohort Description

Our cohort comprised 8,063 chronic pain patients with 3,485,348 accompanying notes (see Table 1 for demographic data). We identified 150,270 total regular expression pattern matches based on the ABC criteria. 52 patients (0.64%) had no associated notes based on search criteria. 1,329 patients (16.5%) had an SUD ICD code on least 2 days while 714 patients (8.9%) had OUD ICD code on at least 2 days.

**Table 1.** Descriptive statistics of demographic characteristics.

| Variable | Mean (SD) | Median (IQR) |
| --- | --- | --- |
| Age at Earliest Chronic Pain Diagnosis (years) | 56.2 (16.3) | 58.0 (46.3-68.5) |
| EHR Duration (years) | 2.5 (2.8) | 1.6 (0.3-3.9) |
| Gender (n = 8,063) | **No. (%)** |  |
| - Female | 5081 (63.0) |  |
| - Male | 2982 (37.0) |  |
| Ethnicity (n = 8,063) |  |  |
| - Hispanic/Latino | 135 (1.7) |  |
| - Non-Hispanic/Latino | 7898 (98.0) |  |
| - Unknown | 30 (0.4) |  |
| Race (n = 8,063) |  |  |
| - Asian | 76 (1.0) |  |
| - Black | 1336 (16.6) |  |
| - Other or more than 1 Race | 122 (1.5) |  |
| - Unknown | 30 (0.4) |  |
| - White | 6499 (80.6) |  |

### ABC Item Performance

Ninety-nine out of 100 patients in the hold-out test set had clinical notes that met inclusion criteria. Of the 20 ABC items, 15 items were associated with positive matches in the test set (see Table 2). When evaluating each regular expression-based ABC item against the test set, sensitivity ranged 0.00-0.90, specificity ranged 0.43-1.00, positive predictive value ranged 0.00-1.00, negative predictive value ranged 0.49-0.60, and F1-scores ranged 0.00-0.73. The ABC item with the highest F1-score performance was “Patient used illicit drugs or evidences problem drinking. The proportion of patients with an ABC item match was similar between the entire cohort and the Test Set with one exception. The “Patient reports minimal/inadequate relief from narcotic analgesic” item had a greater proportion of matches in the Test Set than the entire cohort. The item-level pairwise phi correlation coefficients from the ABC instrument yielded values between –0.01 through 0.32, indicating low item-level correlation in the entire dataset. The sensitivity (recall) and precision (positive predictive value) of the total ABC score (as compared to manual review) were higher than that of the ICD codes (see Figure 1). The total ABC score achieved F1 values of 0.74 (total score >= 1), 0.73 (total score >= 2), 0.56 (total score >= 3), and an AUC of 0.82 (see Figure 2) compared to the manual review, all of which were equal to or better than the ICD-based system (see below).

**Figure 1.**
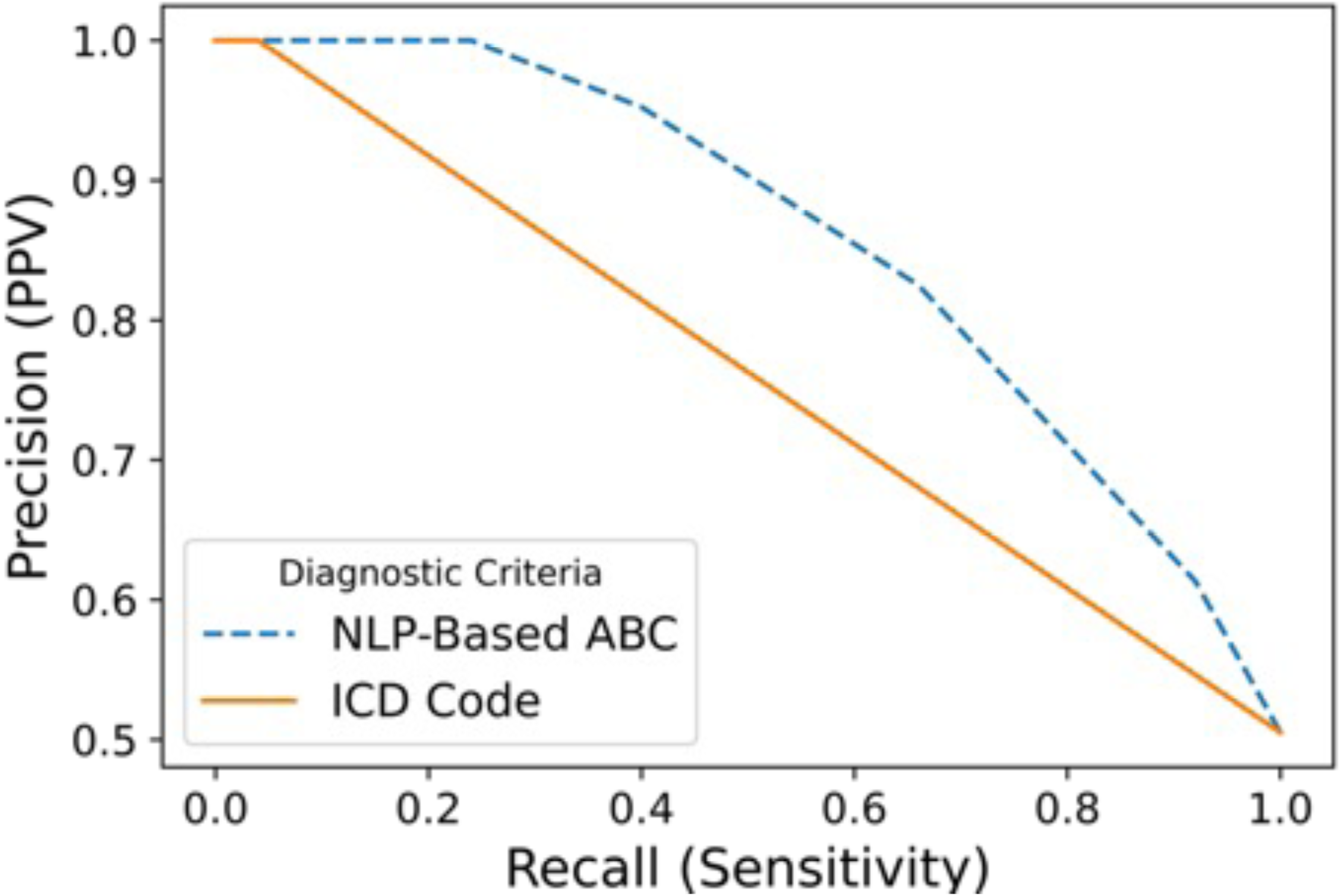
Recall-precision curve of combined score from regular expression-based ABC instrument compared to manual review (blue) and ICD codes (orange).

**Figure 2.**
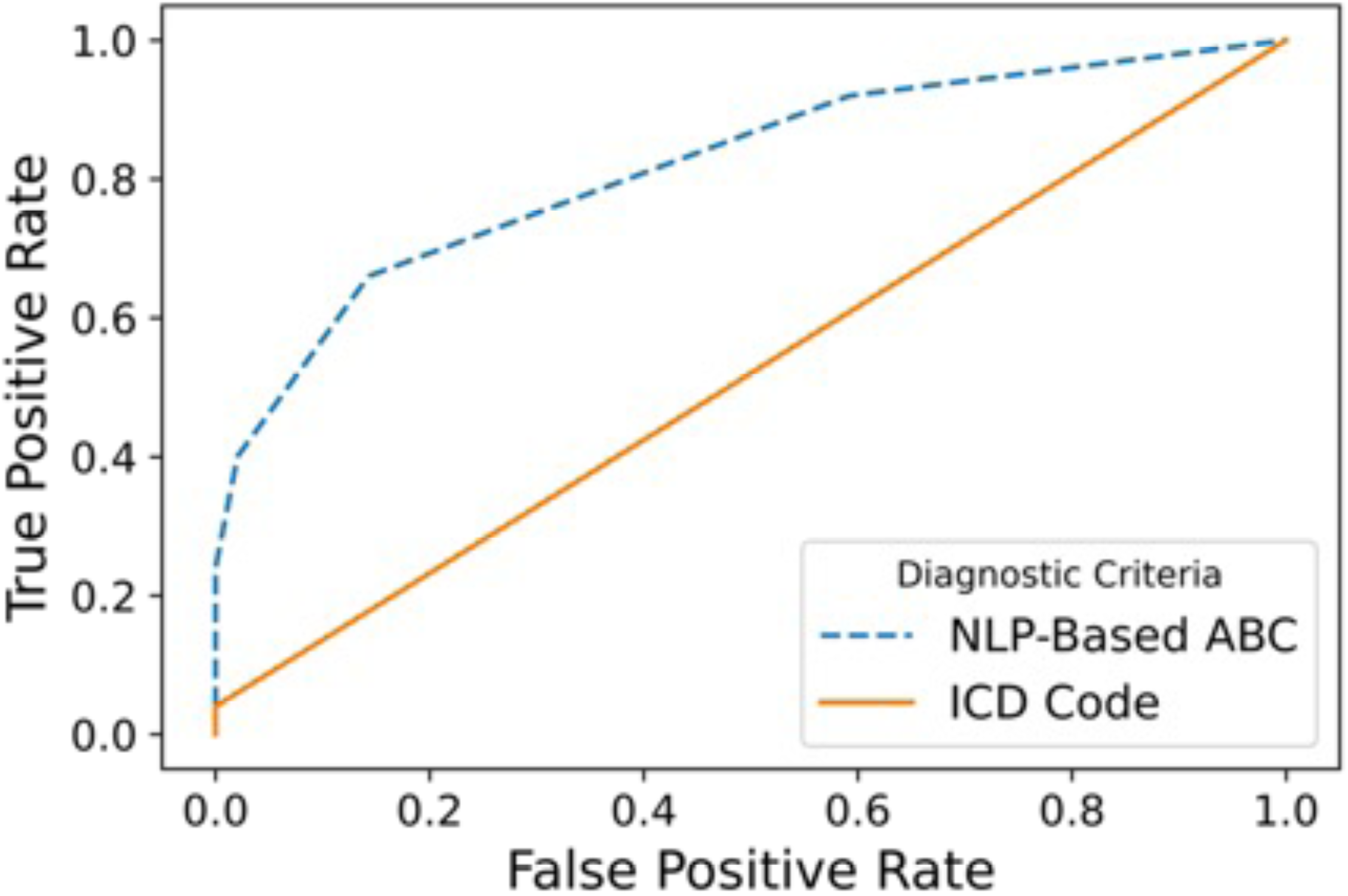
Area under the receiver operating characteristic curve of ABC instrument compared to manual review (blue) and ICD codes (orange).

**Table 2.** Characteristics of each regular expression-based ABC item, ranked in descending order of F1-scores.

| ABC Item | No. (%) of Patients with an ABC Item Match |  | Performance in Test Set |  |  |  |  |
| --- | --- | --- | --- | --- | --- | --- | --- |
|  | Entire Cohort (n = 8063) | Test Set (n = 99) | Sensitivity | Specificity | PPV | NPV | F1 - Score |
| Discussion of analgesic meds was the predominant issue of visit | 6171 (76.5) | 73 (73.7) | 0.90 | 0.43 | 0.62 | 0.81 | 0.73 |
| Patient used illicit drugs or evidences problem drinking | 1454 (18.0) | 21 (21.2) | 0.38 | 0.96 | 0.90 | 0.60 | 0.54 |
| Patient reports minimal/inadequate relief from narcotic analgesic | 1025 (12.7) | 20 (20.2) | 0.34 | 0.94 | 0.85 | 0.58 | 0.49 |
| <sup>a</sup> Patient appears sedated or confuses (e.g., slurred speech, unresponsive) | 1003 (12.4) | 17 (17.2) | 0.26 | 0.92 | 0.76 | 0.55 | 0.39 |
| Patient expressed a strong preference for a specific type of analgesic or a specific route of administration | 1022 (12.7) | 10 (10.1) | 0.20 | 1.00 | 1.00 | 0.55 | 0.33 |
| Patient used analgesics PRN when prescription is for time contingent use | 251 (3.1) | 5 (5.1) | 0.10 | 1.00 | 1.00 | 0.52 | 0.18 |
| Patient received narcotics from more than one provider | 231 (2.9) | 4 (4.0) | 0.08 | 1.00 | 1.00 | 0.52 | 0.15 |

**Table 2. Characteristics of each regular expression-based ABC item, ranked in descending order of F1-scores. (continued)**
|  | **No. (%) of Patients with an ABC Item Match (continued)** |  | **Performance in Test Set (continued)** |  |  |  |  |
| --- | --- | --- | --- | --- | --- | --- | --- |
|  | Entire Cohort (n = 8063) | Test Set (n = 99) | Sensitivity | Specificity | PPV | NPV | F1 - Score |
| **ABC Item (continued)** |  |  |  |  |  |  |  |
| Patient expresses concern about future availability of narcotic | 241 (3.0) | 3 (3.0) | 0.06 | 1.00 | 1.00 | 0.51 | 0.11 |
| Patient exhibited lack of interest in rehab or self-management | 117 (1.5) | 3 (3.0) | 0.06 | 1.00 | 1.00 | 0.51 | 0.11 |
| Patient ran out of meds early | 168 (2.1) | 2 (2.0) | 0.04 | 1.00 | 1.00 | 0.51 | 0.08 |
| Patient used more narcotic than prescribed | 163 (2.0) | 1 (1.0) | 0.02 | 1.00 | 1.00 | 0.50 | 0.04 |
| Significant others express concern over patient's use of analgesics | 137 (1.7) | 1 (1.0) | 0.02 | 1.00 | 1.00 | 0.50 | 0.04 |
| Patient has hoarded meds. | 130 (1.6) | 1 (1.0) | 0.02 | 1.00 | 1.00 | 0.50 | 0.04 |
| Patient has increased use of narcotic | 124 (1.5) | 1 (1.0) | 0.02 | 1.00 | 1.00 | 0.50 | 0.04 |
| Patient expresses worries about addiction | 18 (0.2) | 1 (1.0) | 0.02 | 1.00 | 1.00 | 0.50 | 0.04 |
| <sup>b</sup> Patient indicated she or he "needs" or "must have" analgesic meds | 66 (0.8) | 0 (0.0) | 0.00 | 1.00 | 0.00 | 0.49 | NA |
| Patient indicated difficulty with using medication agreement | 62 (0.8) | 0 (0.0) | 0.00 | 1.00 | 0.00 | 0.49 | NA |
| Patient bought meds on the streets | 31 (0.4) | 0 (0.0) | 0.00 | 1.00 | 0.00 | 0.49 | NA |
| Patient reports worsened relationships with family | 16 (0.2) | 0 (0.0) | 0.00 | 1.00 | 0.00 | 0.49 | NA |
| Patient misrepresented analgesic prescription or use | 2 (0.0) | 0 (0.0) | 0.00 | 1.00 | 0.00 | 0.49 | NA |

<sup>a</sup> Future work could focus on excluding mentions near other sedating interventions (e.g., patient-controlled analgesia in the hospital setting). <sup>b</sup> Regular expressions were unable to capture the intent of this item.

### ICD Code Performance

The prevalence of OUD ICD codes (on at least 2 separate days) among individuals with some or high evidence of OUD was small (3.0% and 5.9%, respectively; see eTable 1). The prevalence of ICD codes for the more generic condition of substance use disorder was higher than that of OUD (15.2% and 47.1%, respectively; see eTable 1). Figure 1 illustrates the sensitivity (recall) versus precision (PPV) of the OUD ICD codes as compared to the manual review and were lower than that of the ABC score. The OUD ICD score achieved an F1 value of 0.08 and an AUC of 0.52 (see Figure 2) compared to the manual review. As the total ABC score increased, the proportion of individuals with an OUD ICD code on at least 2 separate days increased (see Table 3).

**Table 3.** Prevalence of OUD ICD codes among patients in the entire cohort, stratified by total ABC score.

| **ABC Score** | **Sample Size** | **At least 2 days of ICD Code Present for OUD** |
| --- | --- | --- |
| 0 | 1,702 | 15 (0.9%) |
| 1 | 3,200 | 126 (3.9%) |
| 2 | 1,470 | 182 (12.4%) |
| 3 | 760 | 111 (14.6%) |
| 4 | 375 | 93 (24.8%) |
| 5 | 192 | 71 (37.0%) |
| 6 | 106 | 55 (51.9%) |
| 7 | 46 | 26 (56.5%) |
| 8 | 21 | 9 (42.9%) |
| 9 | 11 | 8 (72.7%) |
| 10 | 11 | 10 (90.9%) |
| 11 | 4 | 3 (75.0%) |
| 12 | 3 | 3 (100.0%) |
| 13 | 1 | 1 (100.0%) |

## Discussion

In this study of patients with chronic pain, we demonstrated the feasibility of using regular expressions (an NLP technique) to automate the ABC instrument for identifying OUD problematic opioid use in clinical notes. To our knowledge, this is the first attempt to automate the ABC instrument. Even with a limited set of the patients’ clinical notes for review, the automated approach identified individuals with problematic opioid use that were missed by ICD codes.

Our finding that ICD codes have limitations in identifying problematic opioid use in EHRs aligns with prior work. Clinical text and administrative billing codes have each been consistently shown to provide unique information that can assist with identification of problematic opioid use.^11,12,16^ The idea that a single domain of the EHR (e.g., ICD codes, laboratory values, clinical notes) can adequately yield a valid phenotype is increasingly gaining scrutiny.^20,22,23^

The ABC instrument has historically been completed by clinicians to assess for risk of aberrant opioid use, with a threshold of 3 indicating potential inappropriate use;^1,24,25^ however, some studies have used a threshold of 2 positive items to identify concerning opioid behaviors.^26,27^ In this study, we found the best performance (based on F1 measures) was a threshold of 1. While a low cut-off score on a 20-item measure might not be intuitive, it could be a reflection of the highly-diagnostic nature of many of the ABC items for opioid misuse (e.g. buying drugs on the street).

In previous research, the most frequently endorsed items of the ABC instrument that have been associated with global clinical judgment include: (a) “difficulty with using medication agreement,” (b) “increased use of narcotics (since last visit),” (c) “used more narcotics than prescribed,” and (d) “patient indicated that s/he ‘needs’ or ‘must have’ analgesic meds^.”1^ In our study, we identified that (a) was associated with no matches in our test set; however, we did identify text matches with (b) and (c), albeit with low F1 scores. We were unable to develop regular expressions that appropriately represented the intent of item (d). This could be due to difficulty in capturing intent using current NLP tools or that some expressions will still necessitate patient self-report. Notably, the best-performing items were “Discussion of analgesic meds was the predominant issue of visit” and “Patient used illicit drugs or evidences problem drinking.” The former finding was surprising to us because regular expressions are not typically helpful in understanding a theme or making a generalization about text. For the implementation of this item, we simply required the mention of at least two opioid terms be near each other. The latter finding is perhaps unsurprising because SUD is highly co-morbid with OUD, and while SUD is also rarely reported, SUD behaviors are more frequently documented in the EHR than OUD behaviors.^28^

Our study also has its limitations. We used a single medical center’s EHR data and in a somewhat homogenous cohort of patients with chronic pain. Keywords determined by OUD subject area experts might not be representative of the variety of language in EHR notes. It is possible additional input from external stakeholders and manual reviews of a larger corpus of notes could generate more expressions that would capture additional examples of representing ABC items in clinical notes, which would also enhance generalizability.

## Conclusion

In this study, we leveraged a publicly-available, valid, and reliable instrument for developing our text-based scoring system. Benefits of this method are interpretability (i.e., one can review examples in the chart that match a regular expression), generalizability to other organizations given it can be implemented in multiple software programs, and outperformance of diagnostic codes. Additionally, NLP approaches can serve as one of many approaches to identifying problematic opioid use. For example, a more complex system that also includes laboratory values, coding data, or any other data elements can be constructed. As the regular expressions are improved with input from other investigators and evaluated in larger sets of clinical notes, there is potential for this approach to both automate EHR note reviews and assist in representing problematic opioid use as a continuum rather than a binary condition. Advances in this area will continue to facilitate earlier identification of people at-risk for, and suffering from, problematic opioid use, which will create new opportunities for studying long-term sequelae of opioid pain management.

## Data Sharing Statement

Although the data are de-identified, our data use agreement prohibits sharing raw text data with external entities. Those who would like to review our aggregated data should contact the corresponding author to request a copy of the aggregated data. All code is publicly available at [unblinded URL after review].

## Data Availability

Although the data are probabilistically de-identified, our data use agreement prohibits sharing raw text data with external entities. Those who would like to review our aggregated data should contact the corresponding author to request a copy of the aggregated data.

## Acknowledgments

Dr. Jeffery had full access to all of the data in the study and takes responsibility for the integrity of the data and the accuracy of the data analysis.

*Concept and design:* All authors

*Acquisition, analysis, or interpretation of data:* All authors

*Drafting of the manuscript:* Chatham, Jeffery

*Critical revision of the manuscript for important intellectual content:* All authors

*Statistical analysis:* Chatham, Bradley, Jeffery

*Obtained funding:* Jeffery

*Administrative, technical, or material support:* Jeffery, Samuels

*Supervision:* Schirle, Sanchez-Roige, Samuels, Jeffery

The authors have no conflicts of interest to declare.

Drs. Jeffery, Samuels, Sanchez-Roige, and Schirle received support from the National Institute on Drug Abuse (NIDA) under Award Number DP1DA056667. Dr. Jeffery received support for this work from the Agency for Healthcare Research and Quality (AHRQ) and the Patient-Centered Outcomes Research Institute (PCORI) under Award Number K12 HS026395. Dr. Sanchez-Roige was supported by funds from the California Tobacco-Related Disease Research Program (TRDRP; Grant Number T29KT0526 & T32IR5226), Dr. Sanchez-Roige was also supported by NIDA DP1DA054394. Dr. Schirle received support for this work from the National Institute of Nursing Research (NINR) under Award Number K2313242701. Mr. Chatham received support for this work from the Vanderbilt University Merit Scholarship Stipend Program.

The content is solely the responsibility of the authors and does not necessarily represent the official views of AHRQ, PCORI, the United States Government, or the National Institutes of Health.

