## Supplemental Materials for "Detecting Problematic Opioid Use in the Electronic Health Record: Automation of the Addiction Behaviors Checklist in a Chronic Pain Population"

**Supplemental Content**

1. **Appendix A: Full List of ICD Codes**
2. **Appendix B: List of opioid-related terms**
3. **Appendix C: List of negation-related terms**
4. **Regular expressions**
5. **eTable 1**

**Appendix A**

**Full list of ICD codes**

Chronic Pain

ICD-9-CM: '338.2', '338.21', '338.22', '338.28', '338.29'

ICD-10-CM: 'G89.2', 'G89.21', 'G89.22', 'G89.28', 'G89.29'

Opioid Use Disorder

ICD-9-CM: '304', '304.0', '304.00', '304.01', '304.02', '304.03', '304.7', '304.70', '304.71', '304.72', '304.73', '305.5', '305.50', '305.51', '305.52', '305.53'

ICD-10-CM: 'F11','F11.1','F11.10', 'F11.120','F11.122','F11.129', 'F11.14', 'F11.150','F11.151','F11.159', 'F11.181','F11.182','F11.188', 'F11.19', 'F11.2','F11.20','F11.21','F11.220','F11.221','F11.222','F11.229', 'F11.23','F11.24','F11.250','F11.251','F11.259','F11.281','F11.282','F11.288','F11.29'

ICD-10: 'F11.0','F11.1','F11.2','F11.3','F11.4','F11.5','F11.7','F11.9'

Substance Use Disorder

ICD-9-CM: '292', '292.0', '292.1', '292.11', '292.12', '292.2', '292.8', '292.82', '292.83', '292.84', '292.85', '292.89', '292.9', '304', '304.0', '304.00', '304.01', '304.02', '304.03', '304.1', '304.10', '304.11', '304.12', '304.13', '304.2', '304.20', '304.21', '304.22', '304.23', '304.3', '304.30', '304.31', '304.32', '304.33', '304.4', '304.40', '304.41', '304.42', '304.43', '304.5', '304.50', '304.51', '304.52', '304.53', '304.6', '304.60', '304.61', '304.62', '304.63', '304.7', '304.70', '304.71', '304.72', '304.73', '304.8', '304.80', '304.81', '304.82', '304.83', '304.9', '304.90', '304.91', '304.92', '304.93', '305', '305.2', '305.20', '305.21', '305.22', '305.23', '305.3', '305.30', '305.31', '305.32', '305.33', '305.4', '305.40', '305.41', '305.42', '305.43', '305.5', '305.50', '305.51', '305.52', '305.53', '305.6', '305.60', '305.61', '305.62', '305.63', '305.7', '305.70', '305.71', '305.72', '305.73', '305.8', '305.80', '305.81', '305.82', '305.83', '305.9', '305.90', '305.91', '305.92', '305.93', '306', '357.6', '648.3', '648.30', '648.31', '648.32', '648.33', '648.34', '965.00', '965.01', '965.02'

ICD-10-CM: 'F11','F11.1','F11.10','F11.120','F11.122','F11.129','F11.14','F11.150','F11.151','F11.159','F11.181','F11.182','F11.188','F11.19','F11.2','F11.20','F11.21','F11.220','F11.221','F11.222','F11.229','F11.23','F11.24','F11.250','F11.251','F11.259','F11.281','F11.282','F11.288','F11.29','F11.90','F11.920','F11.922','F11.929','F11.93','F11.94','F11.950','F11.951','F11.959','F11.981','F11.982','F11.988','F11.99','F12','F12.1','F12.10','F12.120','F12.122','F12.129','F12.150','F12.151','F12.159','F12.180','F12.188','F12.19','F12.2','F12.20','F12.21','F12.220','F12.221','F12.222','F12.229','F12.250','F12.251','F12.259','F12.280','F12.288','F12.29','F12.90','F12.920','F12.922','F12.929','F12.950','F12.951','F12.959','F12.980','F12.988','F12.99','F13','F13.10','F13.120','F13.129','F13.14','F13.150','F13.151','F13.159','F13.180','F13.181','F13.182','F13.188','F13.19','F13.20','F13.21','F13.220','F13.221','F13.229','F13.230','F13.231','F13.232','F13.239','F13.24','F13.250','F13.251','F13.259','F13.26','F13.27','F13.280','F13.281','F13.282','F13.288','F13.29','F13.90','F13.920','F13.929','F13.930','F13.931','F13.932','F13.939','F13.94','F13.950','F13.951','F13.959','F13.96','F13.97','F13.980','F13.981','F13.982','F13.988','F13.99','F14','F14.1','F14.10','F14.120','F14.122','F14.129','F14.14','F14.150','F14.151','F14.159','F14.180','F14.181','F14.182','F14.188','F14.19','F14.2','F14.20','F14.21','F14.220','F14.221','F14.222','F14.229','F14.23','F14.24','F14.250','F14.251','F14.259','F14.280','F14.281','F14.282','F14.288','F14.29','F14.90','F14.920','F14.922','F14.929','F14.94','F14.950','F14.951','F14.959','F14.980','F14.981','F14.982','F14.988','F14.99','F15','F15.10','F15.120','F15.122','F15.129','F15.14','F15.150','F15.151','F15.159','F15.180','F15.181','F15.182','F15.188','F15.19','F15.20','F15.21','F15.220','F15.221','F15.222','F15.229','F15.23','F15.24','F15.250','F15.251','F15.259','F15.280','F15.281','F15.282','F15.288','F15.29','F15.90','F15.920','F15.922','F15.929','F15.93','F15.94','F15.950','F15.951','F15.959','F15.980','F15.981','F15.982','F15.988','F15.99','F16','F16.1','F16.10','F16.120','F16.122','F16.129','F16.14','F16.150','F16.151','F16.159','F16.180','F16.183','F16.188','F16.19','F16.2','F16.20','F16.21','F16.220','F16.221','F16.229','F16.24','F16.250','F16.251','F16.259','F16.280','F16.283','F16.288','F16.29','F16.90','F16.920','F16.929','F16.94','F16.950','F16.951','F16.959','F16.980','F16.983','F16.988','F16.99','F17.203','F17.208','F17.209','F17.213','F17.218','F17.219','F17.223','F17.228','F17.229','F17.293','F17.298','F17.299','F18','F18.1','F18.10','F18.120','F18.129','F18.14','F18.150','F18.151','F18.159','F18.17','F18.180','F18.188','F18.19','F18.2','F18.20','F18.21','F18.220','F18.221','F18.229','F18.24','F18.250','F18.251','F18.259','F18.27','F18.280','F18.288','F18.29','F18.90','F18.920','F18.929','F18.94','F18.950','F18.951','F18.959','F18.97','F18.980','F18.988','F18.99','F19.10','F19.120','F19.122','F19.129','F19.14','F19.150','F19.151','F19.159','F19.16','F19.17','F19.180','F19.181','F19.182','F19.188','F19.19','F19.20','F19.21','F19.220','F19.221','F19.222','F19.229','F19.230','F19.231','F19.232','F19.239','F19.24','F19.250','F19.251','F19.259','F19.26','F19.27','F19.280','F19.281','F19.282','F19.288','F19.29','F19.90','F19.920','F19.922','F19.929','F19.930','F19.931','F19.932','F19.939','F19.94','F19.950','F19.951','F19.959','F19.96','F19.97','F19.980','F19.981','F19.982','F19.988','F19.99','F55','F55.0','F55.1','F55.2','F55.3','F55.4','F55.8','O99.32','O99.320','O99.321','O99.322','O99.323','O99.324','O99.325'

ICD-10: 'F10','F11.0','F11.1','F11.2','F11.3','F11.4','F11.5','F11.7','F11.9','F12.0','F12.1','F12.2','F12.5','F12.8','F12.9','F13.0','F13.1','F13.2','F13.3','F13.4','F14.0','F14.1','F14.2','F14.5','F14.9','F15.0','F15.1','F15.2','F15.3','F15.5','F15.8','F15.9','F16.1','F16.2','F16.3','F16.5','F16.8','F16.9','F55','T40.1','T40.3'

**Appendix B**

**List of opioid-related terms**

'pain med', 'opioid', 'opiod', '\bnarc', 'analges', 'suboxone', 'Avinza', 'codeine', 'dilaudid', 'fentanyl', 'hydrocodone', 'morphine', 'opana', 'opiate', 'oxycodone', 'oxycontin', 'oxymorphone', 'percocet', 'roxicodone', 'sufentanyl', 'vicodin', 'lortab', 'hydromorphone', 'abstral', 'actiq', 'alfentanil', 'arymo', 'ascomp', 'astramorph', 'avinza', 'belbuca', 'brompheniramine', 'bunavail', 'buprenex', 'buprenorphine', 'butalbital', 'butorphanol', 'butrans', 'capcof', 'carisoprodol', 'cheratussin', 'coditussin', 'conzip', 'demerol', 'dexbrompheniramine', 'dihydrocodeine', 'diskets', 'dolophine', 'durmorph', 'embeda', 'endacof', 'endocet', 'exalgo', 'fentora', 'fioricet', 'flowtuss', 'guaifenesin', 'histex', 'hycet', 'hycofenix', 'hydrocodone', 'hydromorphone', 'hysingla', 'ibudone', 'infumorph', 'iophen', 'iorinal', 'kadian', 'lazanda', 'levorphanol', 'lorcet', 'lotruss', 'meperidine', 'methadone', 'methadose', 'morphabond', 'morphine', 'ms contin', 'nalbuphine', 'nalocet', 'ninjacof', 'nucynta', 'obredon', 'opana', 'opium', 'orco', 'oxaydo', 'oxecta', 'oxycodone', 'panlor', 'paregoric', 'pentazocine', 'percocet', 'phenylhistine', 'primlev', 'pro-clear', 'probuphine', 'promethazine', 'psuedoephedrine', 'relcof', 'remifentanil', 'reprexain', 'rezira', 'robafen', 'roxicodone', 'rydex', 'suboxone', 'subsys', 'sufentanil', 'synalgos', 'talwin', 'tapentado', 'tramadol', 'trezix', 'triplidine', 'trymine', 'tusnel', 'tussicpas', 'ultiva', 'ultracet', 'ultram', 'verdrocet', 'vicodin', 'vicoprofen', 'virtussin', 'xartemix', 'xodol', 'xtampza', 'zamicet', 'zodryl', 'zubsolv', 'zutripro', 'zylon'

**Appendix C**

**List of negation-related terms**

'\bno\b', '\bnot\b', 'denies', 'denial', 'doubt', 'never', 'negative for'

**Appendix D**

**Regular expressions**

checklist = {'1a' : {'lab' : 'Since last visit: #1 "Patient used illicit drugs or evidences problem drinking" #1a Illicit drugs',
 'pat' : r'((illicit drug)|mariju|cocai|heroin|polysubst|methamphetamine|amphetamine|\becstasy|(street drugs)|IVDU|(IV drug))',
 'col_name' : 'illicit_drugs',
 'opioid' : True,
 'negation' : True,
 'preview' : True},
 '1b' : {'lab' : '#1b Problematic alcohol use',
 'pat' : r'((problem)([^\.]{1,10})(drink|alcoh|etoh))|(((drink|alcoh|etoh))([^\.]{1,10})(abuse|addic|ism|depend)(?![^\.]{1,10}(father|mother)))',
 'col_name' : 'problem_drinking',
 'opioid' : False,
 'negation' : True,
 'preview' : True,
 'common_fp' : ['mother', 'sister', 'father', 'brother', 'aunt', 'uncle']},
 '1c' : {'lab' : '1c. DUIs',
 'pat' : r'(\bDUI\b)',
 'col_name' : 'dui',
 'opioid' : False,
 'negation' : True,
 'preview' : True},
 '2' : {'lab' : '#2 "Patient has hoarded meds"',
 'pat' : r'(\bhoard|stash|(left over)|\bstoring|stockpil)',
 'col_name' : 'hoarding',
 'opioid' : True,
 'negation' : True,
 'preview' : True,
 'common_fp' : ['hoarder']},
 '3' : {'lab' : '#3 "Patient used more narcotic than prescribed"',
 'pat' : r'((more+(?=.{1,50}than presc))|(ta?o?o?ke?i?n?g? extra(?![^\.]{1,10}strength)))',
 'col_name' : 'more_narcotic',
 'opioid' : True,
 'negation' : True,
 'preview' : True},
 '4' : {'lab' : '#4 "Patient ran out of meds early"',
 'pat' : r'(((running|ran) \bout\b(?=.{1,50}earl))|((\bout\b(?=.{1,50}earl)))|(too early to refill)|(refill early))',
 'col_name' : 'ran_out_early',
 'opioid' : True,
 'negation' : True,
 'preview' : True,
 'common_fp' : ['out of bed']},
 '5a' : {'lab' : '#5a "Patient has increased use of narcotics" 5a. Increased narcotic/opioid use',
 'pat' : r'((increased|increasing) use)',
 'col_name' : 'increased_op',
 'opioid' : True,
 'negation' : True,
 'preview' : True},
 '5b' : {'lab' : '#5b Dose escalation (alternative to increased use)',
 'pat' : r'((escalate?d?i?o?n?(?=((.{1,10})dos)))|(dose?a?g?e?(?=((.{1,10})escalat))))',
 'col_name' : 'escalation_op',
 'opioid' : True,
 'negation' : True,
 'preview' : True},
 '6' : {'lab' : '#6 "Patient used analgesics PRN when prescription is for time contingent use"',
 'pat' : r'(not?n? ?-?adher)|(non?t?-? ?compli)',
 'col_name' : 'prn',
 'opioid' : True,
 'negation' : False,
 'preview' : True,
 'common_fp' : ['mepitel nonadherent dressing', 'diet', 'regimen']},
 '7a' : {'lab' : '#7 "Patient received narcotics from more than one provider"',
 'pat' : r'(more than one|multiple)(?=((.{1,50}(provider|doctor|pharmac|prescrib))))',
 'col_name' : 'multiple_sources_x',
 'opioid' : True,
 'negation' : False,
 'preview' : True,
 'common_fp' : ['diagnoses']},
 '7b' : {'lab' : 'Alternate "More than one provider" Search',
 'pat' : r'(more than one|multiple)(?=((.{1,50}(provider|doctor|pharmac|prescrib))))',
 'col_name' : 'multiple_sources_y',
 'opioid' : True,
 'negation' : True,
 'preview' : True,
 'common_fp' : ['diagnoses']},
 '8' : {'lab' : '#8 "Patient bought meds on the streets"', #alternate: r'((med|drug).{1,10}(on the street))|((on the street).{1,10}(med|drug))'
 'pat' : r'(on the street)', #original: 28 matches, alternate: 4 matches
 'col_name' : 'bought_on_street',
 'opioid' : True,
 'negation' : False,
 'preview' : True},
 '9' : {'lab' : '#9 "Patient appears sedated or confused (e.g., slurred speech, unresponsive)"',
 'pat' : r'(sedated|confused|(slurred speech)|unresponsive)',
 'col_name' : 'sedated',
 'opioid' : True,
 'negation' : False,
 'preview' : True},
 '10' : {'lab' : '#10 "Patient expresses worries about addiction"',
 'pat' : r'(worr.{1,25}addic)',
 'col_name' : 'worries_about_addiction',
 'opioid' : False,
 'negation' : True,
 'preview' : True},
 '11a' : {'lab' : '#11 "Patient expressed a strong preference for a specific type of analgesic or a specific route of administration"',
 'pat' : r'((patient|pt) (request|prefer|(preferr?s? to take)))',
 'col_name' : 'strong_preference',
 'opioid' : True,
 'negation' : True,
 'preview' : True,
 'common_fp' : ['pharm']},
 '11b' : {'lab' : '#11b Strong Preference, IV',
 'pat' : 'r((patient|pt) (request|prefer|(prefers to take))([^\.]{1,10}IV))',
 'col_name' : 'strong_preference_IV',
 'opioid' : False,
 'negation' : False,
 'preview' : True},
 '12a' : {'lab' : '#12a "Patient expresses concern about future availability of narcotic"', #change take to tak and add opioid detection
 'pat' : r'(((take)[^\.]{1,25})(away))',
 'col_name' : 'future_availability_1',
 'opioid' : True,
 'negation' : False,
 'preview' : True,
 'common_fp' : ['disp', 'rfl', 'allerg']},
 '12b' : {'lab' : '#12b Future Availability: Loss of medication',
 'pat' : r'((((won|can)\'?t)[^\.]{1,25}(get))|(loss of medication))',
 'col_name' : 'future_availability_2',
 'opioid' : True,
 'negation' : False,
 'preview' : True},
 '13' : {'lab' : '#13 "Patient reports worsened relationships with family"',
 'pat' : r'(family (dynamics|problem))|(relationship problem)|((lost|(took))[^\.]{1,25}(kids|children|\bson|daughter))',
 'col_name' : 'worsened_relationships',
 'opioid' : True,
 'negation' : False,
 'preview' : True},
 '14' : {'lab' : '#14 "Patient misrepresented analgesic prescription or use"', #opioid detection, add \b in front of lie
 'pat' : r'(misrepresent|(lied? about)|(\blying about))',
 'col_name' : 'misrep_use',
 'opioid' : True,
 'negation' : False,
 'preview' : True},
 '15' : {'lab' : '#15 "Patient indicated she or he ‚Äò‚Äòneeds‚Äô‚Äô or ‚Äò‚Äòmust have‚Äô‚Äô analgesic meds"',
 'pat' : '((must have)|demands)',
 'col_name' : 'needs_must_have' ,
 'opioid' : True,
 'negation' : False,
 'preview' : True},
 '16' : {'lab' : '#16 "Discussion of analgesic meds was the predominant issue of visit"',
 'pat' : r'(pain med)|opioid|opiod| narc|analges|suboxone|Avinza|codeine|dilaudid|fentanyl|hydrocodone|morphine|opana|opiate|oxycodone|oxycontin|oxymorphone|percocet|roxicodone|sufentanyl|vicodin|lortab|hydromorphone|abstral|actiq|alfentanil|arymo|ascomp|astramorph|avinza|belbuca|brompheniramine|bunavail|buprenex|buprenorphine|butalbital|butorphanol|butrans|capcof|carisoprodol|cheratussin|coditussin|conzip|demerol|dexbrompheniramine|dihydrocodeine|diskets|dolophine|durmorph|embeda|endacof|endocet|exalgo|fentora|fioricet|flowtuss|guaifenesin|histex|hycet|hycofenix|hydrocodone|hydromorphone|hysingla|ibudone|infumorph|iophen|iorinal|kadian|lazanda|levorphanol|lorcet|lotruss|meperidine|methadone|methadose|morphabond|morphine|ms contin|nalbuphine|nalocet|ninjacof|nucynta|obredon|opana|opium|orco|oxaydo|oxecta|oxycodone|panlor|paregoric|pentazocine|percocet|phenylhistine|primlev|pro-clear|probuphine|promethazine|psuedoephedrine|relcof|remifentanil|reprexain|rezira|robafen|roxicodone|rydex|suboxone|subsys|sufentanil|synalgos|talwin|tapentado|tramadol|trezix|triplidine|trymine|tusnel|tussicpas|ultiva|ultracet|ultram|verdrocet|vicodin|vicoprofen|virtussin|xartemix|xodol|xtampza|zamicet|zodryl|zubsolv|zutripro|zylon',
 'col_name' : 'opioid_tag',
 'opioid' : True,
 'negation' : True,
 'preview' : True,
 'common_fp' : ['disp', 'rfl', 'allerg', 'mg']},
 '17' : {'lab' : '#17 "Patient exhibited lack of interest in rehab or self-management"',
 'pat' : r'((not|no[^\.(need)]{1,25}(interest|seen|want|go))|resistant|dislike|refuse)([^\.]{1,25})(rehab|(pain management)|(pain clinic))',
 'col_name' : 'lack_interest_rehab',
 'opioid' : True,
 'negation' : True,
 'preview' : True},
 '18a' : {'lab' : '#18a "Patient reports minimal/inadequate relief from narcotic analgesic"',
 'pat' : r'(not?\b|minimal|limited)([^\.]{1,25})(relief (from|with|w/t|))',
 'col_name' : 'minimal_relief_x',
 'opioid' : True,
 'negation' : True,
 'preview' : True},
 '18b' : {'lab' : 'Patient reports minimal/inadequate relief: Tolerance',
 'pat' : r'(\btoleran)',
 'col_name' : 'tolerance',
 'opioid' : True,
 'negation' : True,
 'preview' : True,
 'common_fp' : ['activity']},
 '19' : {'lab' : '#19 "Patient indicated difficulty with using medication agreement"',
 'pat' : r'((medication|med|opioi?d|narcotic) (agreement|contract) (breach|violat|problem|issue|fail))|((violat|problem|issue|fail|breach)[^\.]{1,25}(medication|med|opioi?d|narcotic) (agreement|contract))',
 'col_name' : 'med_agreement',
 'opioid' : False,
 'negation' : False,
 'preview' : True},
 '20' : {'lab' : '#20 (Other) "Significant others express concern over patient‚Äôs use of analgesics"',
 'pat' : r'(wife|\bmother|father|husband|daughter|aunt|uncle|\bson|(significant other)|(family member))([^\.]{1,25})(concern|worr)',
 'col_name' : 'SO_concern',
 'opioid' : True,
 'negation' : False,
 'preview' : True},
 }

**eTable 1. Prevalence of OUD and SUD ICD codes among patients in the Test Set, stratified by degree of OUD evidence on manual review.**

|  | **At least 2 days of ICD Code Present for OUD (n=2)** | **At least 2 days of ICD Code Present for SUD (n=14)** |
| --- | --- | --- |
| No evidence (n=49) | 0 (0.0%) | 1 (2.0%) |
| Some evidence (n=33) | 1 (3.0%) | 5 (15.2%) |
| High evidence (n=17) | 1 (5.9%) | 8 (47.1%) |
